# Comparative Safety of Therapies for Hyperthyroidism Planning Pregnancy: A Network Meta-analysis

**DOI:** 10.1101/2020.05.11.20097774

**Authors:** Xiaohua Fang, Huibin Huang, Wei Lin, Jixing Liang, Junping Wen, Gang Chen

**Author notes:** Correspondence and reprints request: Gang Chen Gang Chen, MD, PhD, Department of Endocrinology, Shengli Clinical Medical College of Fujian Medical University, Fujian Provincial Hospital, Fuzhou, 350001, China.

## Abstract

**Objective:** Our intention was to compare the pregnancy safety in women with hyperthyroidism seeking future pregnancy after ATD, RAI or thyroidectomy treatment and to explore the optimum treatment strategy. We hope the results will guide the choice for endocrinologists and patients, and be beneficial for promoting eugenics.

**Methods:** We searched multiple databases though December 2019. The outcome indicators were abortion, preterm birth, IUGR and birth defect. We conducted a frequency-framework network meta-analysis by using Stata and R software. The results of the intervention comparison were expressed as OR with 95%CI, and ranking was assessed using surface under the cumulative ranking (SUCRA) probabilities.

**Results:** The network meta-analysis included 4 retrospective cohort studies with a total enrollment of 480 patients. ①Thyroidectomy had a lower risk of abortion than RAI [OR=0.77, 95%CI (0.23, 2.56)] and ATD [OR=0.68, 95%CI (0.21, 2.21)]. RAI had a lower risk of abortion than ATD [OR=0.88, 95%CI (0.45, 1.75)]. Based on SUCRA results, thyroidectomy (0.698) was followed by RAI (0.494), ATD (0.308). Thyroidectomy (60.7%) had the highest probability of being first compared to RAI (27.0%) and ATD (12.4%). ②Thyroidectomy had a lower risk of preterm birth than RAI [OR=0.80, 95%CI (0.26, 2.44)] and ATD [OR=0.79, 95%CI (0.39, 1.59)]. RAI had a lower risk of preterm birth than ATD [OR=0.98, 95%CI (0.42, 2.33)]. Based on SUCRA results, thyroidectomy (0.703) was followed by RAI (0.430), ATD (0.367). Thyroidectomy (57.8%) had the highest probability of being first compared with RAI (30.5%) and ATD (11.8%). ③Thyroidectomy had a lower risk of IUGR than RAI [OR=0.28, 95%CI (0.03, 3.02)] and ATD [OR=0.83, 95%CI (0.14, 4.86)]. RAI had a higher risk of IUGR than ATD [OR=3.02, 95%CI (0.60, 15.27)]. Based on SUCRA results, thyroidectomy (0.717) was followed by ATD (0.663), RAI (0.120). Thyroidectomy (56.6%) had the highest probability of being first compared with RAI (5.4%) and ATD (37.9%). ④Thyroidectomy had a lower risk of birth defect than RAI [OR=0.70, 95%CI (0.02, 30.34)] and ATD [OR=0.23, 95%CI (0.01, 4.52)]. RAI had a lower risk of birth defect than ATD [OR=0.32, 95%CI (0.03, 3.12)] (Table 2). Based on SUCRA results, thyroidectomy (0.70) was followed by RAI (0.629), ATD (0.171). Thyroidectomy (55.9%) had the highest probability of being first compared with RAI (41.3%) and ATD (2.8%).

**Conclusion:** Thyroidectomy was the optimum option for women with hyperthyroidism seeking near future pregnancy. The future research direction is to include more samples to conduct head-to-head randomized controlled trials or prospective cohort studies, establish inclusion criteria for various pre-pregnancy conditions or further subgroup analysis, and develop more acceptable, safer, and more manageable treatments that allow for the remission of both thyroid function and autoimmune abnormalities.

## Introduction

Hyperthyroidism is common in women of childbearing age^[1]^. The main treatment options for them are antithyroid drugs (ATD), radioactive iodine (RAI) and thyroidectomy. The pregnant safety after different therapies for hyperthyroidism in a patient planning pregnancy is of great concern. However, the relevant guidelines of the Chinese Medical Association Endocrine Society, the European Thyroid Association (ETA) and the American Thyroid Association (ATA) all hold that each therapeutic option carries advantages and disadvantages^[2 3]^, so it is not clear which is the safer option. The early documented surveys showed that clinical endocrinologists from different countries had significantly different clinical practice patterns in the management of a hyperthyroid woman planning pregnancy over the next 6-12 months (the index case). Thyroidectomy is more frequently suggested in Italy than in the EU and USA (Italy: thyroidectomy 54.3%, ATD 36%, RAI 9.7%; EU: ATD 59.3%, thyroidectomy 30.3%, RAI 10.4%; USA: ATD 49.9%, RAI 29.8%, thyroidectomy 20.3%)^[4-6]^. Furthermore, to date, there are no head-to-head trials, and there are few or no pairwise comparative study answering the question. Therefore, in this study, we aimed to do a network meta-analysis to inform clinical practice by comparing the risk of adverse pregnancy outcomes including abortion, premature delivery, intrauterine growth restriction (IUGR) and birth defect.

## Materials and Methods

### Study eligibility

We involved women of childbearing age (20-40y) who were conformed hyperthyroidism and were pregnant after treatment for hyperthyroidism. There were no restrictions on the single, twin or multiple pregnancies. The time interval from treatment to pregnancy was also not limited, except treatment with RAI should be delayed at least 6 months. We did not restrict whether the patient reached a stable euthyroid state before conception. The publications should compare treatments with at least two of the three interventions: ATD (MMI, CMZ, PTU), RAI and thyroidectomy. ATD dose, ATD treatment duration, RAI dose, number of RAI treatments, the extent of thyroidectomy was not restricted. The outcomes were the rate of abortion, premature delivery, IUGR and birth defect. Studies were included in this study were randomized clinical trials (RCT_S_) and observational cohort studies. There was no language and country restrictions.

### Data sources and search strategies

We searched PubMed, Cochrane Library, Embase and CNKI databases from database inception to December 2019. We searched MeSH terms “hyperthyroidism”, “antithyroid agents”, “Methimazole”, “Carbimazole”, “Propylthiouracil”, “radiotherapy”, “thyroidectomy”, “pregnancy”, and “female” with the corresponding free words.

### Study selection and data extraction

Two investigators independently selected eligible studies according to titles, abstracts, and full texts, and extracted the relevant information from the included studies. A form was used to extract the following information: author, data of publication, countries, publication type, the data collection time, sample size, type of treatment, dose and treatment duration of ATD, dose and numbers of RAI, the extent of thyroidectomy (total or subtotal thyroidectomy), the time interval from treatment to pregnancy, mean age, type of hyperthyroidism, disease course, thyroid function and TSH receptor antibodies (TRAb) levels before and during conception, different drugs used during pregnancy, follow-up duration, the number of patients with outcomes.

### Quality assessment

Two investigators independently assessed the risk of bias. The Cochrane collaboration’s risk of bias assessment tool was used for RCTs, and the Newcastle-Ottawa scale (NOS) was used for cohort studies.

## Data analysis

We used Stata (version 16) and R (version 3.5.1) software to do a frequency-framework network meta-analysis. We assessed the heterogeneity for all comparisons using the I^2^ statistic. Consistency between direct and indirect sources of evidence was assessed globally and locally in the closed loops. If at least ten studies were included, we used the funnel plot and Egger’s test to analysis publication bias. The effect estimates were expressed as odds ratio (OR) with 95% confidence interval (95%CI), and the ranking of treatments was assessed using surface under the cumulative ranking (SUCRA) probabilities.

## Results

We identified 6 eligible articles, and 2 articles were excluded because the author couldn’t be contacted for complete data^[7, 8]^. Overall, 4 cohort studies published between 2014 and 2018 were included in the meta-analysis: 3 English articles were from PubMed database^[9-11]^ and 1 Chinese article was from CNKI database^[12]^ (Figure1). These studies involved a total of 480 patients (234 in the ATD group, 147 in the RAI group, 99 in the surgical group). In the surgical group, there were 78% had been performed with subtotal thyroidectomy and 22% had been performed with total thyroidectomy prior to pregnancy. About mean age of the included patients, in the ATD group, 75% of them were 29 years old, and 25% were 26.8±3.5 years old; in the RAI group, 88% of them were 29 years old, and 12% were 31 years old; in the surgery group, 29% of patients were 29 years old, 59% were 26.2±3.4 years old, and 12% were 28 years old. In terms of the mean time interval from treatment to pregnancy, 54% of patients in the ATD group were at least 6 months apart, and 46% were not reported; in the RAI group, 88% of patients had an interval of at least 6 months, and 12% were 32 months (rang, 0.5-96.2 months) apart; in the surgery group, 29% of patients were 3 years (rang, 0-9 years) apart, 12% were 19.6 months (rang, 3-113.1 months) apart, and 59% did not report. In the ATD group, 56% of the patients took ATD or L-T4 during pregnancy, 20% stopped taking medicine, and 24% was not reported; in the RAI group, 82% of the patients took PTU or L-T4 during pregnancy, 18% stopped taking the medicine; in all the surgical group, drugs used during pregnancy were not reported. In regards to thyroid function during pregnancy, in the ATD group, 54% of the included patients had stable thyroid function, and 46% were not reported; in the RAI group, 88% of the patients had stable thyroid function; in the surgical group, 12% of the patients had stable thyroid function, and 56% did not report it. Mean follow-up duration was not clearly indicated. The 4 included articles all reported abortion rate, 3 articles reported premature delivery rate, 2 articles reported IUGR rate, and 2 articles reported birth defect rate. The studies were conducted in the following countries: China, 2; New Zealand, 1; France, 1(Table 1). The methodological quality of included studies saw the Table 2.

**Figure 1.**
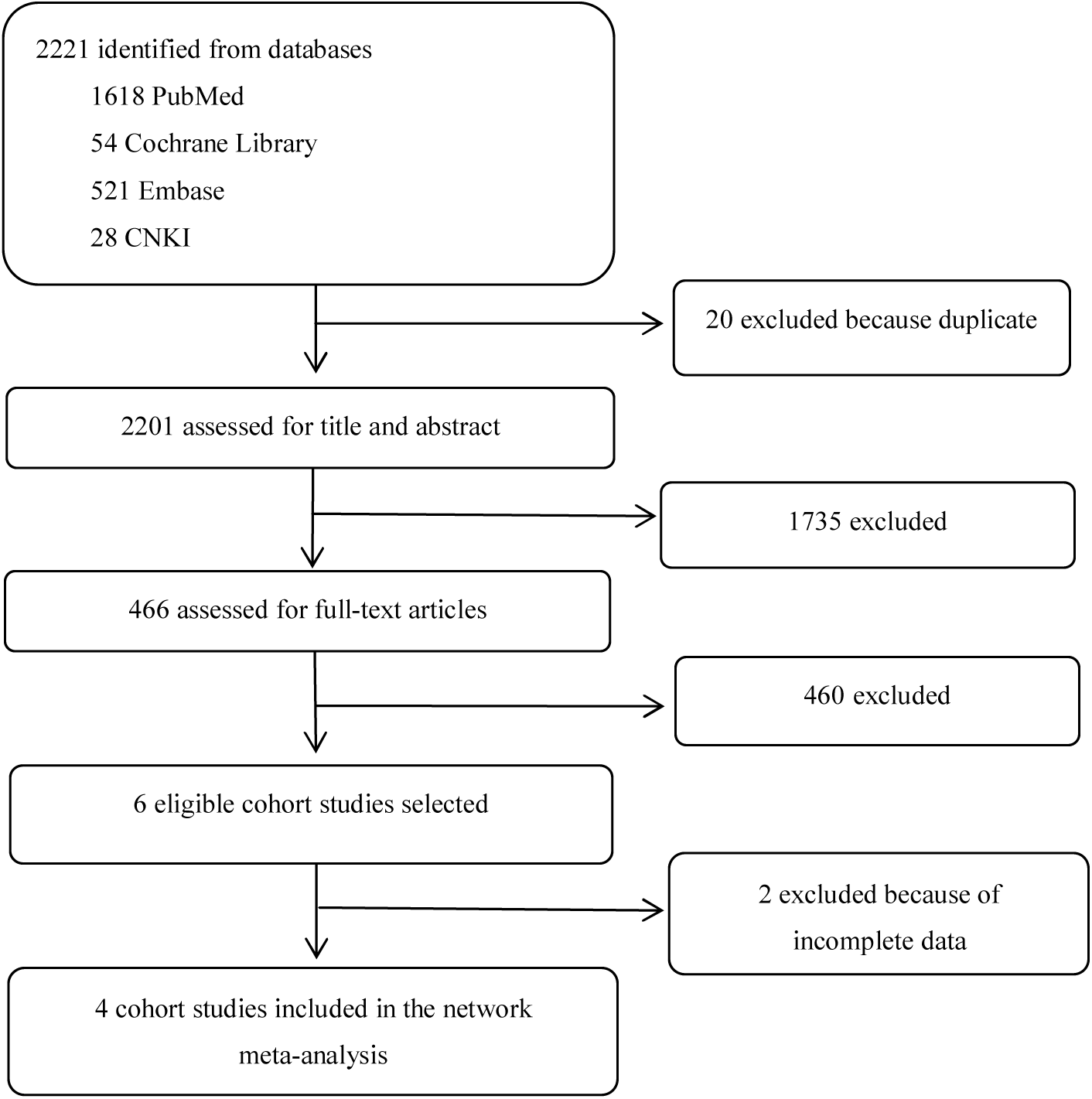
Study selection process

**Table 1.** characteristics of the included studies

| Study name | Country | Diagnosis | Treatment setting | Treatment details | Sample size | Mean age (year) |
| --- | --- | --- | --- | --- | --- | --- |
| Hawken 2016 | France | GD | ATD<br>vs<br>surgery | ATD : Unclear<br>Surgery:TT/ST (12/17) | 49/29 | 29 |
| Zhang 2016 | China | GD | RAI<br>Vs<br>ATD | RAI : 1 /2 /3times<br>( 125/3/2 )<br>ATD : Unclear | 130/127 | 29 |
| Elston 2014 | New Zealand | GD | surgery<br>vs<br>RAI | surgery: TT/ST ( 10/2 )<br>RAI : Unclear | 12/17 | 28/31 |
| Zhiwen Wu2018 | China | hyperthyroidism | surgery<br>vs<br>ATD | surgery : ST<br>ATD : PTU | 58/58 | 26.2±3.4/26.8±3.5 |

| Study name | Mean interval time between treatment and pregnancy | Medication between pregnancy | Thyroid function between pregnancy | Outcomes |
| --- | --- | --- | --- | --- |
| Hawken 2016 | ATD: Unclear<br>Surgery: 3<br>( 0-9 ) years | ATD : T1 : CMZ/PTU/stop<br>(9/15/25)<br>surgery : Unclear | ATD : Unclear<br>Surgery:<br>TSH > 2.5 ( 23 )<br>TSH < 2.5 ( 6 ) | abortion+premature delivery+ IUGR+ birth defect |
| Zhang 2016 | At least 6 months | RAI: PTU/L-T4/no<br>( 43/69/18 )<br>ATD: PTU 或 L-T4/stop<br>( 106/21 ) | stable | abortion+premature delivery+ IUGR+ birth defect |
| Elston<br>2014 | surgery: 19.6<br>( 3-113.1 ) months | Surgery: Unclear | Euthyroid state :<br>Surgery: ( 13/19 ) | abortion |
|  | RAI: 32<br>(0.5-96.2) months | Surgery: ATD/L-T4/no<br>( 5/15/7 ) | RAI: ( 5/23 ) |  |
|  |  |  | TSH > 4 mU/L |  |
|  |  |  | Surgery: ( 9/19 ) |  |
|  |  |  | RAI: ( 12/24 ) |  |
| Zhiwen<br>WU2018 | Unclear | Unclear | Unclear | abortion+premature<br>delivery |

**TT: total thyroidectomy; ST: subtotal thyroidectomy; T1: first trimester of pregnancy**

**Table 2.**
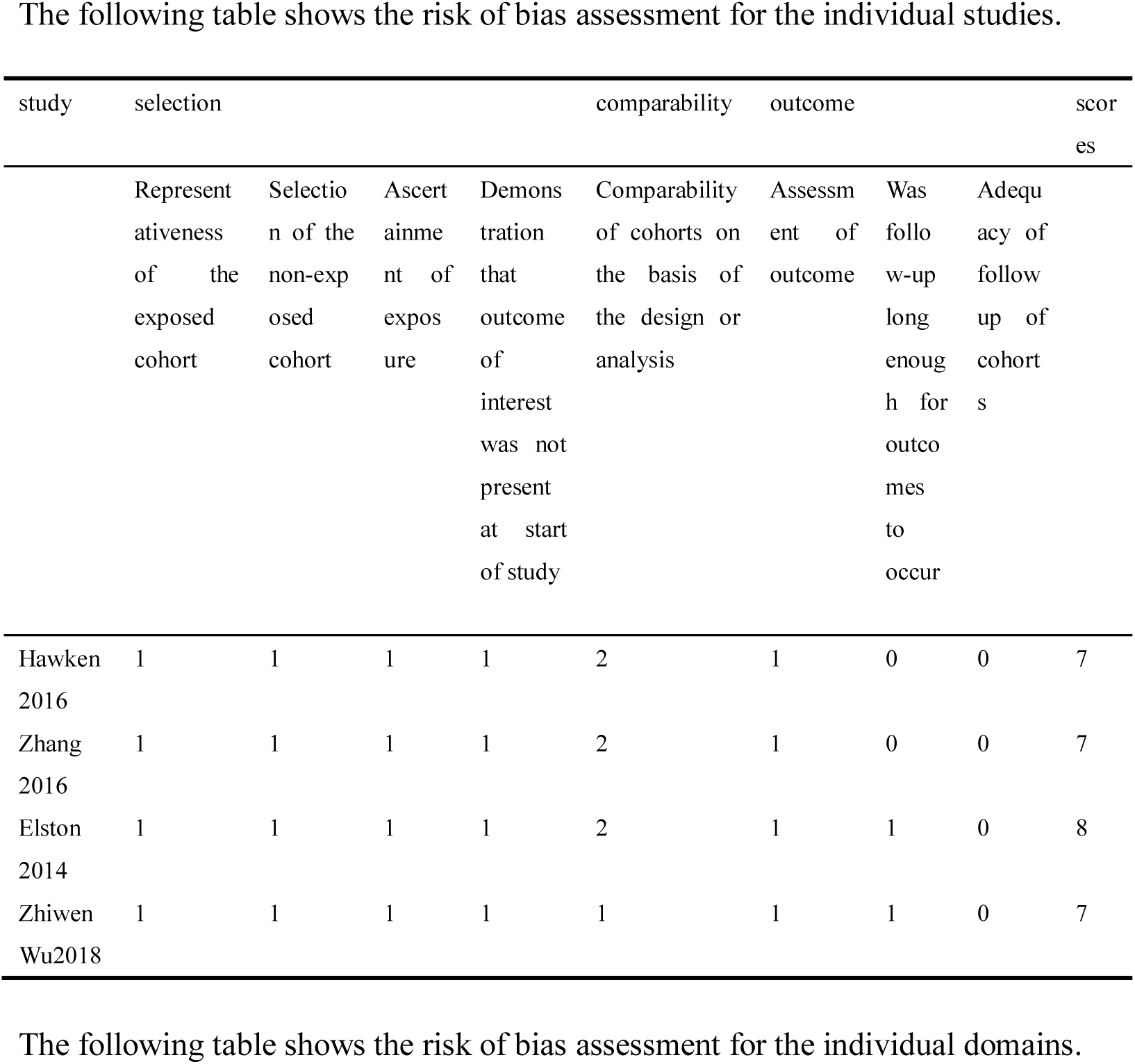
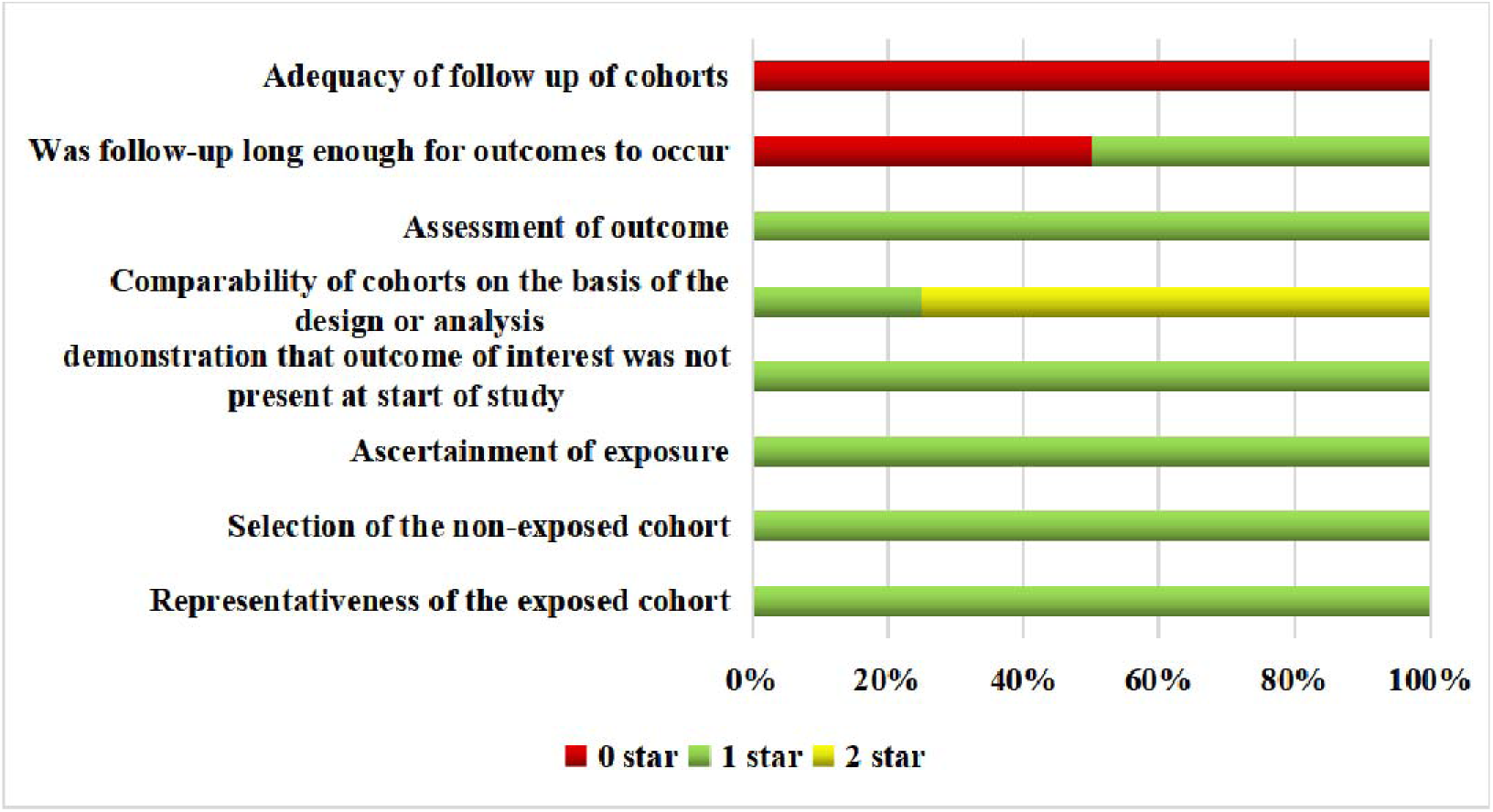
Risk of assessment. The following table shows the risk of bias assessment for the individual studies.

### Network analysis

Figure 2 shows the network plot of pregnancy outcomes including the rate of abortion (A), premature delivery (B), IUGR (C) and birth defect (D). In the analysis, the heterogeneity was all less than 25%, suggesting the low heterogeneity (Table 3). The inconsistency test of abortion wasn’t found statistical difference, showing a good consistency (Figure3, Table 4).

**Figure 2.**
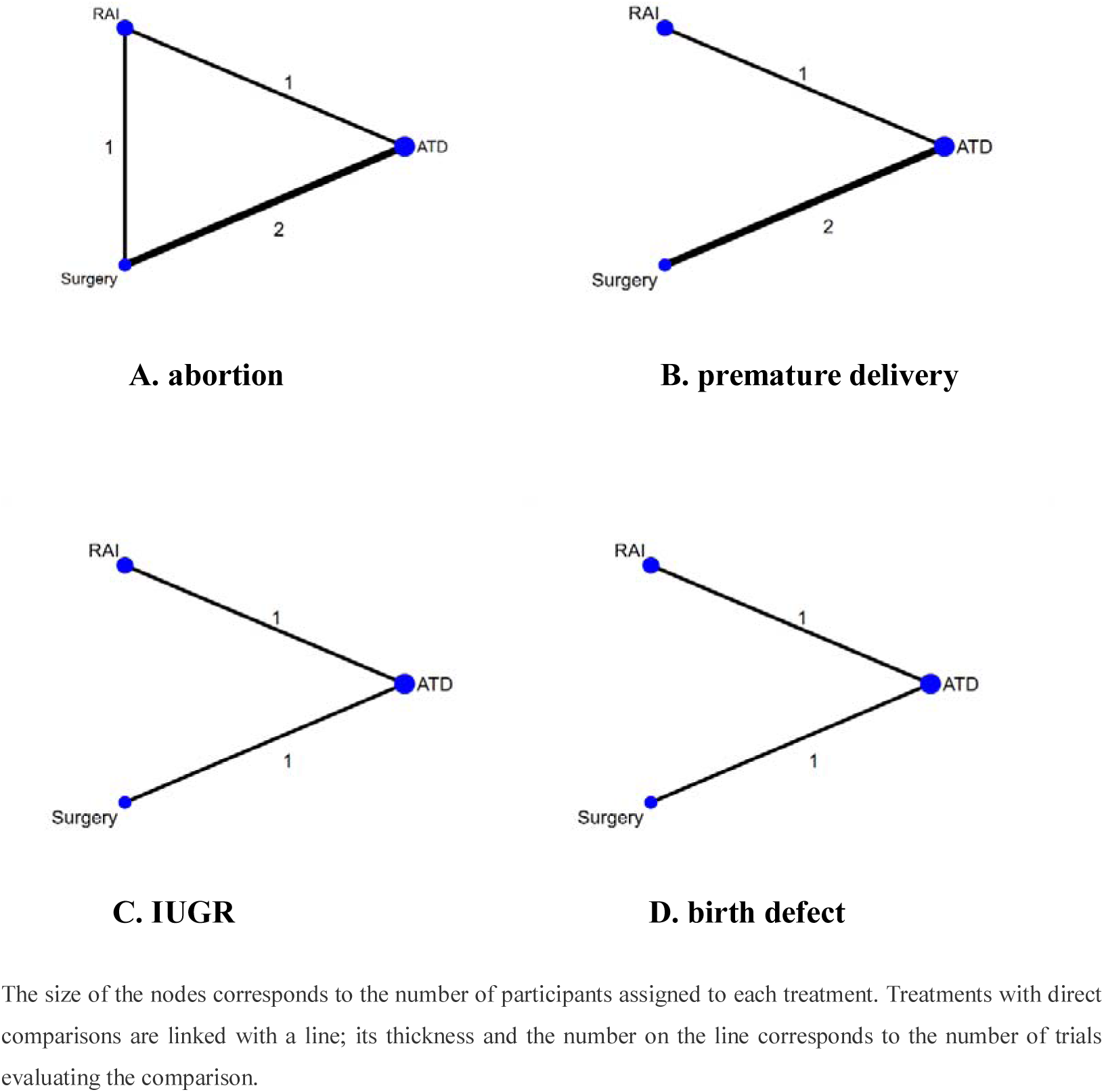
network plot for all the outcomes

**Figure 3.**
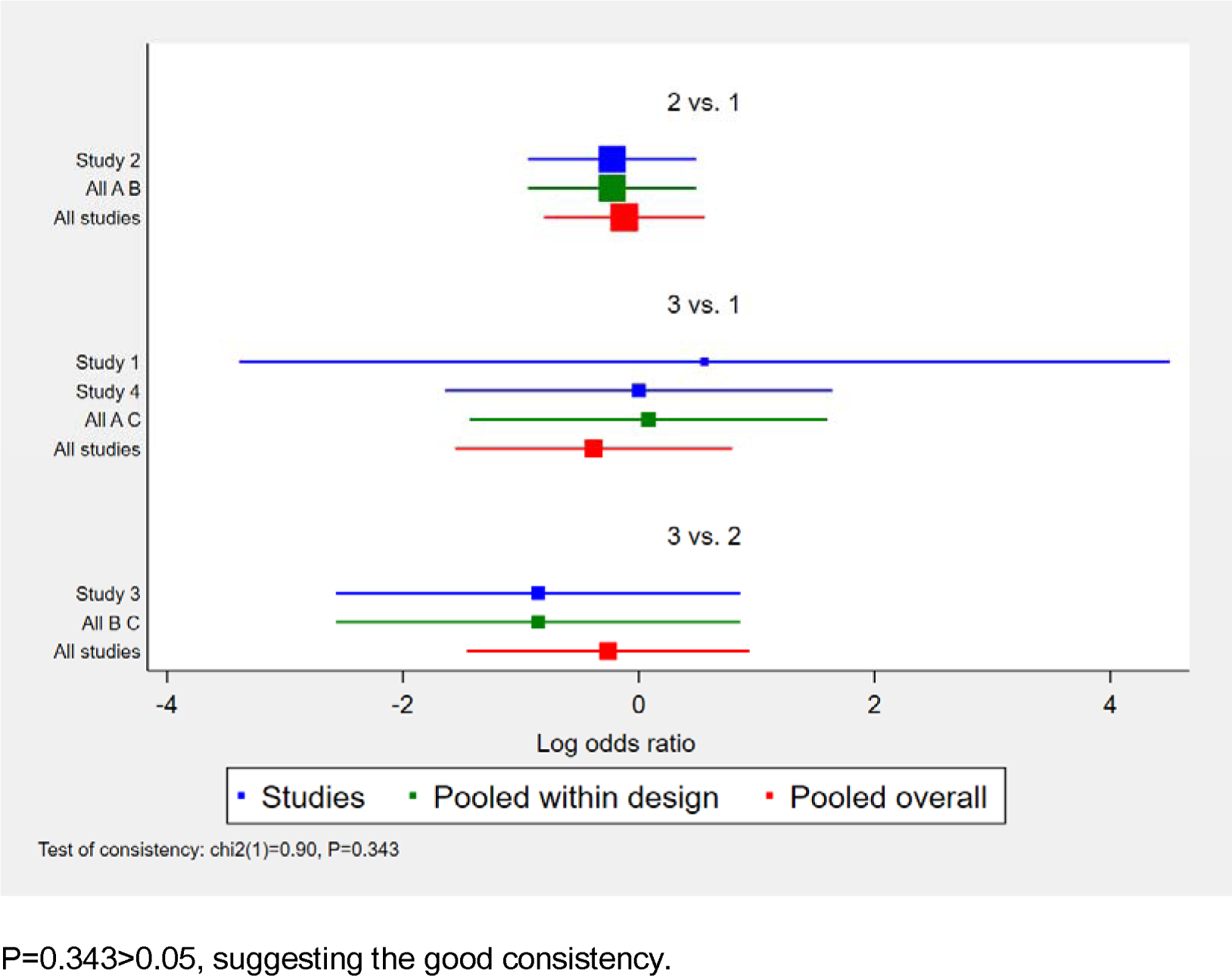
Global inconsistency of abortion

**Table 3.** heterogeneity parameter results in various outcomes

| Outcomes | $I^2$ |
| --- | --- |
| Abortion | 0% |
| Premature delivery | 0% |
| IUGR | 10.4% |
| Birth defect | 0% |

**Table 4.** Local inconsistency of abortion

| Side | Direct |  | Indirect |  | Difference |  |  | tau |
| --- | --- | --- | --- | --- | --- | --- | --- | --- |
|  | coef | Std.Err. | Coef | Std.Err. | Coef | Std.Err. | P>\|z\| |  |
| AB | -.2259175 | .3649147 | .9358467 | 1.168253 | -1.161764 | 1.223919 | 0.343 | 1.68e-07 |
| AC | .0823578 | .7740259 | -1.079407 | .9480839 | 1.161765 | 1.22392 | 0.343 | 3.90e-07 |
| BC | -.8534898 | .8750431 | .3082751 | .8557329 | -1.161765 | 1.22392 | 0.343 | 3.98e-07 |

$P=0.343>0.05$ , suggesting the good consistency.

### Abortion

Thyroidectomy had a lower risk of abortion than RAI [OR=0.77, 95%CI (0.23, 2.56)] and ATD [OR=0.68, 95%CI (0.21, 2.21)]. RAI had a lower risk of abortion than ATD [OR=0.88, 95%CI (0.45, 1.75)] (Table 5). However, the statistical differences in the three treatments were not significant. Based on SUCRA, thyroidectomy (0.698) was ranked first, RAI treatment was second (0.494), ATD was the last (0.308). Thyroidectomy (60.7%) had the highest probability of being first compared to RAI (27.0%) and ATD (12.4%) (Figure 4A, Table 6).

**Figure 4.**
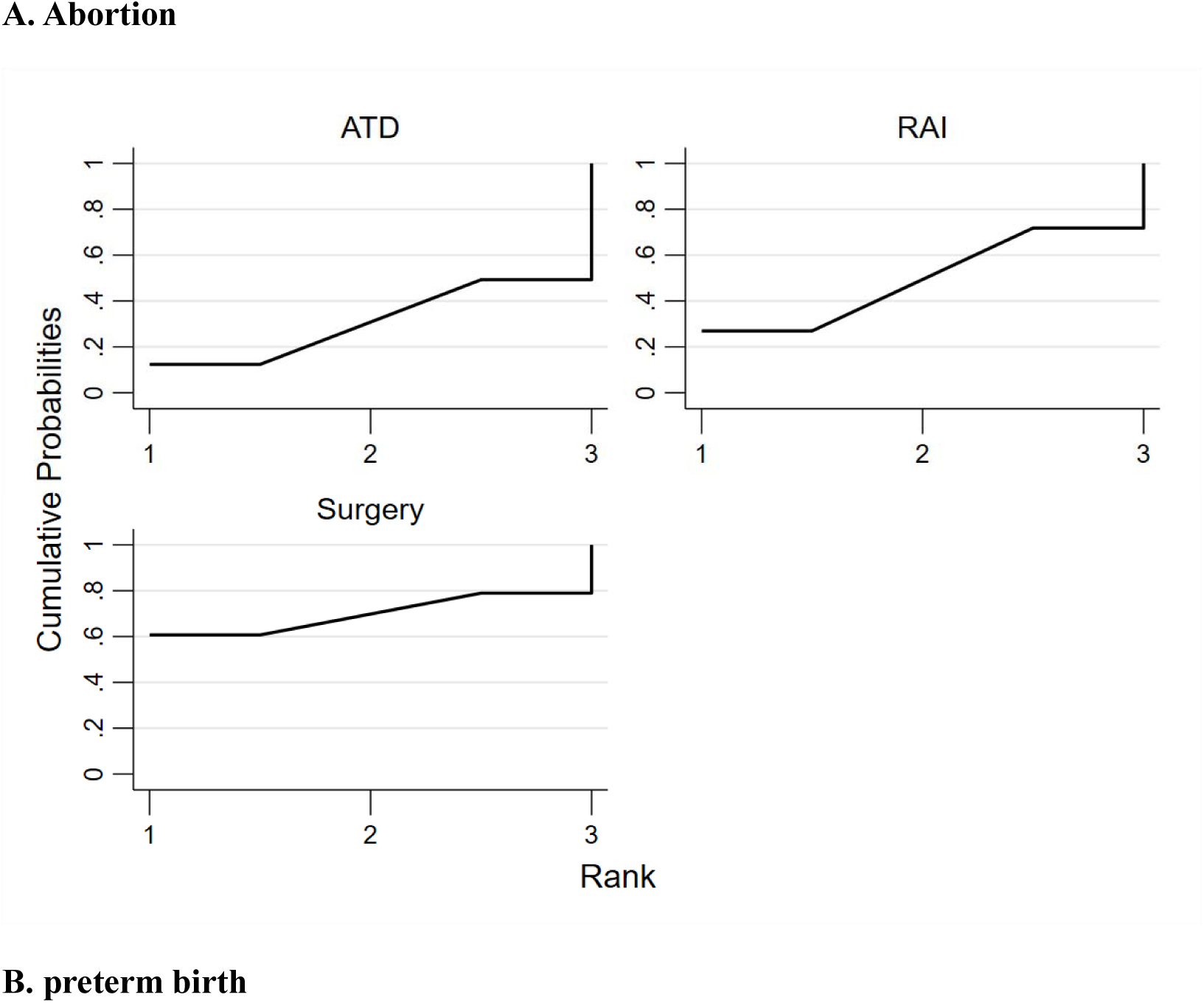
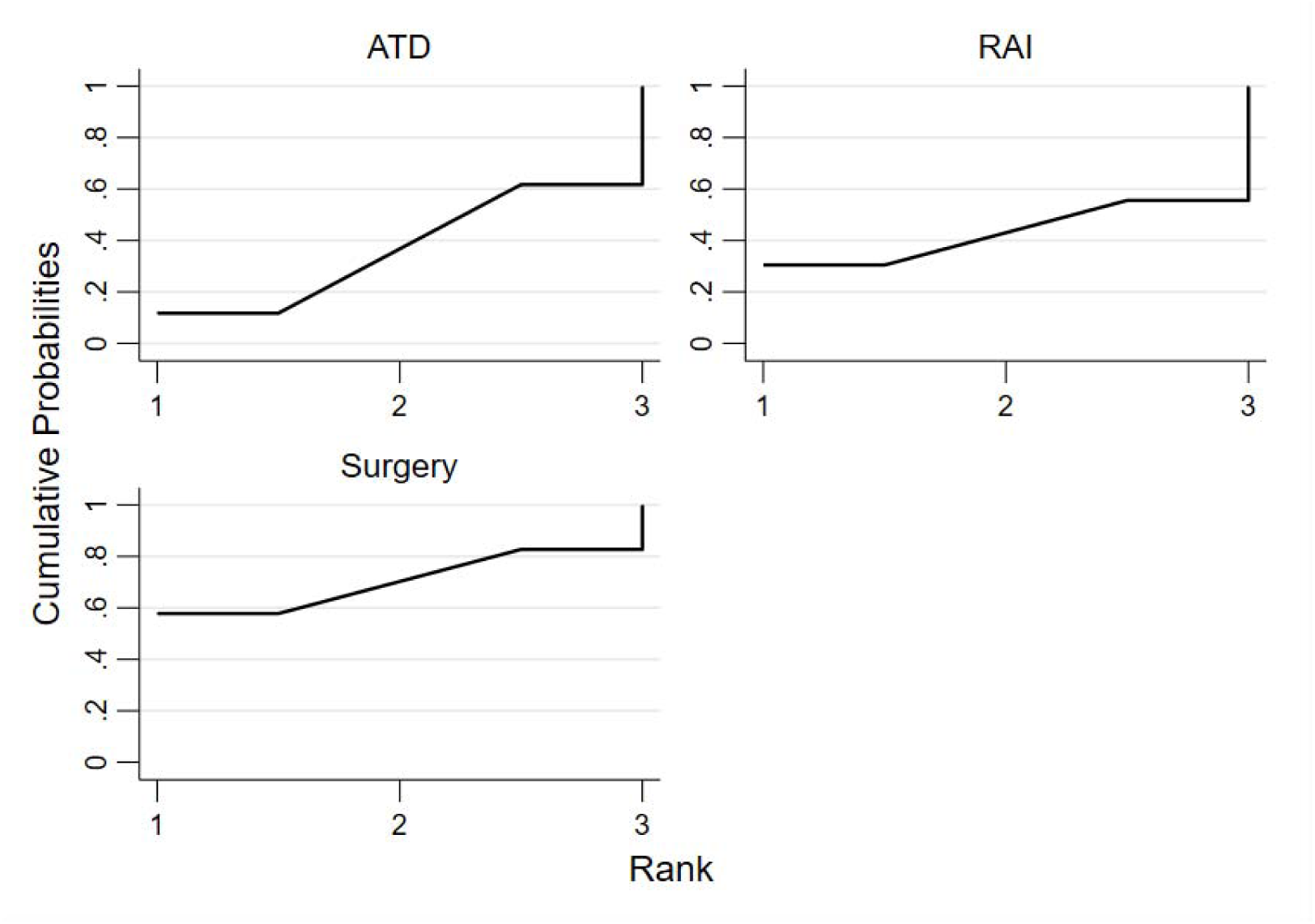
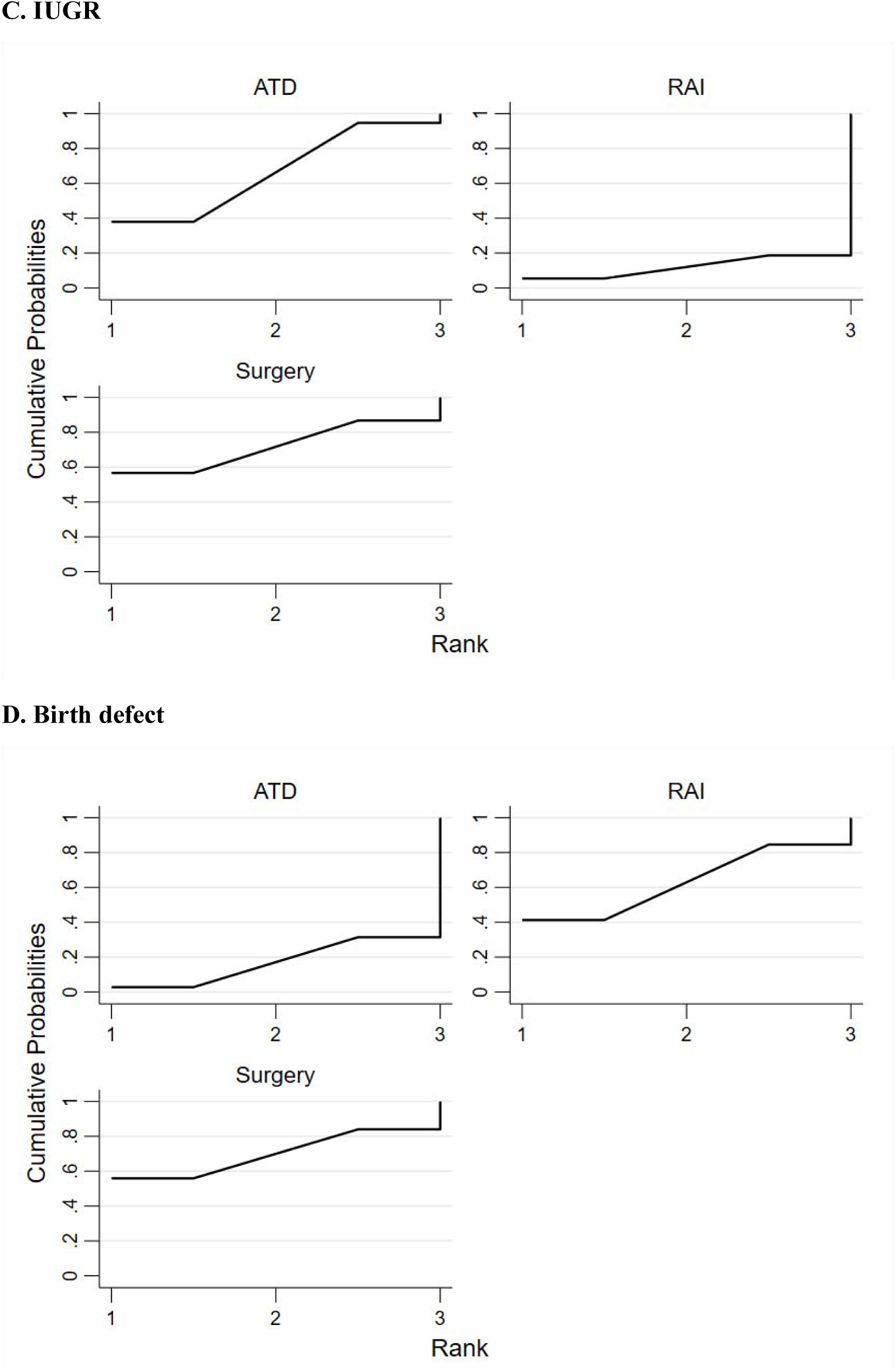
Ranking of comparative pregnancy safety of the interventons

**Table 5.** comparison for pregnancy safety league table

|  |  |  |  |
| --- | --- | --- | --- |
| **Abortion rate** | **ATD** | 0.88<br>(0.45, 1.75) | 0.68<br>(0.21, 2.21) |
|  | 1.13<br>(0.57,2.24) | **RAI** | 0.77<br>(0.23, 2.56) |
|  | 1.47<br>(0.45,4.75) | 1.30<br>(0.39,4.30) | **Surgery** |
| **Preterm birth rate** | **ATD** | 0.98<br>(0.42, 2.33) | 0.79<br>(0.39, 1.59) |
|  | 1.02<br>(0.43,2.41) | **RAI** | 0.80<br>(0.26, 2.44) |
|  | 1.27<br>(0.63,2.55) | 1.24<br>(0.41,3.78) | **Surgery** |
| **IUGR rate** | **ATD** | 3.02<br>(0.60, 15.27) | 0.83<br>(0.14, 4.86) |
|  | 0.33<br>(0.07,1.67) | **RAI** | 0.28<br>(0.03, 3.02) |
|  | 1.20<br>(0.21,7.00) | 3.63<br>(0.33,39.77) | **Surgery** |
| **Birth defect rate** | **ATD** | 0.32<br>(0.03, 3.12) | 0.23<br>(0.01, 4.52) |
|  | 3.12<br>(0.32,30.41) | **RAI** | 0.70<br>(0.02, 30.34) |
|  | 4.44<br>(0.22,89.10) | 1.42<br>(0.03,61.42) | **Surgery** |

**Table 6.** Ranking of comparative pregnancy safety of the interventons

**A. Abortion**
| Treatment | SUCRA | PrBest | MeanRank |
| --- | --- | --- | --- |
| ATD | 30.8 | 12.4 | 02.4 |
| RAI | 49.4 | 27.0 | 02.0 |
| Surgery | 69.8 | 60.7 | 01.6 |

**B. premature delivery**
| Treatment | SUCRA | PrBest | MeanRank |
| --- | --- | --- | --- |
| ATD | 36.7 | 11.8 | 02.3 |
| RAI | 43.0 | 30.5 | 02.1 |
| Surgery | 70.3 | 57.8 | 01.6 |

**C. IUGR**
| Treatment | SUCRA | PrBest | MeanRank |
| --- | --- | --- | --- |
| ATD | 66.3 | 37.9 | 01.7 |
| RAI | 12.0 | 05.4 | 02.8 |
| Surgery | 71.7 | 56.6 | 01.6 |

**D. Birth defect**
| Treatment | SUCRA | PrBest | MeanRank |
| --- | --- | --- | --- |
| ATD | 17.1 | 02.8 | 02.7 |
| RAI | 62.9 | 41.3 | 01.7 |
| Surgery | 70.0 | 55.9 | 01.6 |

### Preterm birth

Thyroidectomy had a lower risk of preterm birth than RAI [OR=0.80, 95%CI (0.26, 2.44)] and ATD [OR=0.79, 95%CI (0.39, 1.59)]. RAI had a lower risk of preterm birth than ATD [OR=0.98, 95%CI (0.42, 2.33)] (Table 5). However, no statistical difference was found among the three groups. Based on SUCRA, thyroidectomy (0.703) was ranked first, RAI was second (0.430), ATD was the last (0.367). Thyroidectomy (57.8%) had the highest probability of being first compared with RAI (30.5%) and ATD (11.8%) (Figure 4B, Table 6).

### IUGR

Thyroidectomy had a lower risk of IUGR than RAI [OR=0.28, 95%CI (0.03, 3.02)] and ATD [OR=0.83, 95%CI (0.14, 4.86)]. RAI had a higher risk of IUGR than ATD [OR=3.02, 95%CI (0.60, 15.27)] (Table 5). However, no statistical difference was found among the three groups. Based on SUCRA, thyroidectomy (0.717) was ranked first, ATD was second (0.663), RAI was the last (0.120). Thyroidectomy (56.6%) had the highest probability of being first compared with RAI (5.4%) and ATD (37.9%) (Figure 4C, Table 6).

### Birth defect

Thyroidectomy had a lower risk of birth defect than RAI *[OR=0*.70, 95*%CI* (0.02, 30.34)] and ATD [*OR*=0.23, 95%*CI* (0.01, 4.52)]. RAI had a lower risk of birth defect than ATD [*OR*=0.32, *95%CI* (0.03, 3.12)] (Table 5). However, no statistical difference was found among the three groups. Based on SUCRA, thyroidectomy (0.70) was ranked first, RAI was second (0.629), ATD was the last (0.171). Thyroidectomy (55.9%) had the highest probability of being first compared with RAI (41.3%) and ATD (2.8%) (Figure 4D, Table 6).

## Discussion

We conducted a network meta-analysis of the pregnancy safety to determine the optimal treatment for hyperthyroidism in a patient planning pregnancy. In the risk of abortion, premature delivery, IUGR and birth defect, we found that thyroidectomy was safer than RAI and ATD, and RAI were safer than ATD except for the IUGR. Assessing the above four adverse pregnancy outcomes comprehensively, the option of thyroidectomy was ranked first for women with hyperthyroidism seeking future pregnancy, RAI was second, and ATD was the last.

According to the available evidence, there may be the following reasons for the last rank of ATD in the above ranking. ATD is a risk factor of birth defect. PTU and MMI/CMZ equally cross the placenta^[13]^. A Denmark national cohort study published in 2013 found that both the MMI/CMZ and PTU were associated with birth defect, but the spectrum of malformations differed^[14]^. Moreover, birth defects could lead to an increase in preterm births. A study between 1995 and 2000 from 13 states showed that birth defects was more than twice among premature babies compared with full-term babies [PR=2.65, 95%CI (2.62, 2.68)]^[15]^. However, a routine shift from MMI/CMZ to PTU in the early pregnancy couldn’t reduce the risk of birth defect (PTU, 8.0%; MMI/CMZ, 9.1%; shift between MMI/CMZ and PTU,10.1%; no ATD, 5.4%; nonexposed, 5.7%; P<0.001)^[14]^. In addition to the increased risk of birth defects, there are problems such as slowly achieving stable euthyroid state, high recurrence rate, long treatment time, and many side effects with ATD therapy. ATD therapy slowly achieving stable euthyroid state leads to women exposed to the risks of pregnancy complicated by hyperthyroidism and the increased need for ATD to control thyroid function during pregnancy. Mr. Hamburger et al. agreed that more aggressive initial therapy (RAI or thyroidectomy) rather than suppressive therapy (ATD) employed before pregnant could prevent pregnancies complicated by hyperthyroidism^[16]^. The reported relapse rate was approximately 50-70%^[2, 17]^. Many meta-analysis had confirmed the high relapse rate of ATD therapy in comparison with RAI and thyroidectomy^[18-20]^. The common adverse effects, including rash, agranulocytosis, hepatic dysfunction, vasculitis, arthritis, were also ascribed to ATD^[21]^. Furthermore, although ATD therapy could lead a fall in TRAb, the effect mainly lasted during the period of ATD administration. After drug withdraw, the abnormal autoimmunity was reactivated in 50% of the patients, resulting in an elevated or reappearance serum TRAb and the relapse of hyperthyroidism^[22]^. Though ATD is an easy oral administration and cheap, it’s not the ideal choice for women with hyperthyroidism seeking future pregnancy.

The reasons why RAI is not the ideal intervention are as follows. TRAb is not easy to relieve after RAI treatment. A 5-year prospective randomized study in Denmark showed that serum TRAb gradually decreased and turned negative after 18 months in about 70-80% of the patients after ATD and thyroidectomy. However, for RAI treatment, the TRAb level deteriorated within 1 year and then began to decline in the following years, and the number of patients with TRAb negative conversion was significantly lower than ATD and thyroidectomy^[22]^. The fetal thyroid gland has the function of secreting thyroid hormone from the 16-17 weeks of pregnancy. Maternal TRAb enters the fetal compartment through the placental barrier, which stimulates the fetal thyroid by binding to the TSH receptor, causing increased thyroid hormone secretion and hyperthyroidism in the fetus and newborn^[23]^. Thyrotoxic fetuses may develop goiter, tachycardia, heart failure, growth retardation, craniosynostosis, increased fetal activity and accelerated bone maturation, leading to premature birth and high early mortality rate^[24, 25]^. In addition, limited information is available on whether TRAb directly contributes to adverse pregnancy outcomes. It was reported that TRAb directly blocked the luteinizing hormone (LH) receptor on the corpus luteum, reducing progesterone secretion^[26]^. Even if the patient’s thyroid function is normal during pregnancy, high levels of TRAb can still lead to adverse pregnancy outcomes. Attention should be paid to monitoring TRAb and fetal growth during pregnancy. Whether RAI therapy for hyperthyroidism will increase genetic damage is also a common concern of many patients and doctors. Although most studies believed that there was no evidence that patients treated with RAI had a significantly increased risk of genetic damage^[27, 28]^. The recurrence of hyperthyroidism after RAI treatment also requires attention. A large retrospective study with over 4 years of follow up showed that nearly 23% of patients relapsed after RAI treatment and needed a second RAI treatment or converted to surgical treatment^[29]^. Laurberg et al. considered the possible teratogenic risks of ATD and the long-lasting TRAb after RAI treatment, suggesting that thyroidectomy was the best treatment option for hyperthyroidism women of childbearing age^[30]^. Although RAI is an easy oral administration and has rapid relief of thyroid function, it does not seem to be the best choice for hyperthyroidism in fertile women.

Davis et al. found that thyroidectomy for hyperthyroidism could restore euthyroid state faster than RAI [3 months (2-7 months) versus 9 months (4-14 months); p < 0.001]^[31]^. A study showed that subtotal thyroidectomy was not an optimal surgical strategy for Graves’ disease (GD) because the recurrence rate (28.7%) was high and the euthyroid rate was lower than total thyroidectomy^[32]^. However, nearly 80% of the included surgical cases underwent subtotal thyroidectomy, and still had a good pregnancy outcome after surgery, probably because most of them had pregnancy before postoperative relapse. Though thyroidectomy is the safer option, it’s not risks free. Surgical complications are a concern. The common complications are transient hypocalcemia and hoarseness, resolving within 6 months in most patients. The most feared complication is permanent recurrent laryngeal nerve injury (0-2%). Other possible complications include a low incidence of superior laryngeal nerve injury, postoperative bleeding, and the risk of anesthesia^[33]^. In addition, after thyroidectomy, a few patients cannot achieve remission of autoimmune abnormality, causing hyperthyroidism in fetus and newborn during pregnancy^[34-36]^. Therefore, women with hyperthyroidism treated with thyroidectomy, need to pay attention to monitoring not only thyroid function to prevent hypothyroidism, but also maternal TRAb and neonatal thyroid function during pregnancy.

Among the 4 outcome indicators, no statistical significance was found, which may be related to the small sample size. Although there was no statistical significance, we cannot ignore its clinical significance. Whether the difference is statistically significant or not, network meta-analysis can always give the optimal intervention measures. Statistics, of course, only gives us a trend, and we need to carefully verify in clinical practice.

This study included the patient with hyperthyroidism who had not achieved euthyroid state before pregnancy because considering the situation of the real-world population and the overall effect with therapies. Although if stricter inclusion criteria were formulated, for example, the ATD group included euthyroid patients before pregnancy, the results of the study would be more targeted. If there are large-scale studies in the future, inclusion criteria can be further limited or subgroup analysis can be feasible. The results of this paper are applicable for women with hyperthyroidism who seek pregnancy in the near future, if not, the results may not be applicable. Because with the extension of the time interval of treatment and pregnancy, the factors affecting pregnancy outcomes become more complex.

We did not identify other network meta-analysis evaluating the comparative safety of therapies for hyperthyroidism in a patient seeking future pregnancy to date. Our results would help clinicians and patients to make choices, and have positive significance for promoting eugenics. Our study has limitations, the primary one being the small number of included studies and enrolled patients, perhaps affecting the stability of our results. Second, our study included cohort studies increasing the risk of inherent bias. Third, network meta-analysis combines direct comparison and indirect comparison of evidence, and the results need careful interpretation. Fourth, because of the limited data, our study didn’t explore the fetal outcome such as cesarean section, macrosomia, fetal distress, fetal death in utero, asphyxia, respiratory distress syndrome, endotracheal intubation, transfer to neonatal intensive care unit (NICU) and death. Fifth, the literature included in the study had short follow-up time about the birth defect, which might underrate the results. Finally, two literatures were not included in this network meta-analysis because the authors were not contacted up to date, which may have an impact on the final results.

Future research direction for therapies for hyperthyroidism in a women seeking near future pregnancy is to include more samples to conduct head-to-head randomized controlled trials or prospective cohort studies, establish inclusion criteria of various pre-pregnancy conditions or further subgroup analysis, and develop more acceptable, safer, and more manageable treatments that allow for the remission of both thyroid function and autoimmune abnormalities.

## Data Availability

All data included in this study are available upon request by contact with the corresponding author.

## References

[1] CarlÉ A, Pedersen I B, Knudsen N, et al. Epidemiology of subtypes of hyperthyroidism in Denmark: a population-based study [J]. European journal of endocrinology, 2011,164(5): 801–809.

[2] Alexander E K, Pearce E N, Brent G A, et al. 2017 Guidelines of the American Thyroid Association for the Diagnosis and Management of Thyroid Disease During Pregnancy and the Postpartum[J]. Thyroid, 2017,27(3): 315–389.

[3] Kahaly G J, Bartalena L, Hegedus L, et al. 2018 European Thyroid Association Guideline for the Management of Graves’ Hyperthyroidism [J]. Eur Thyroid J, 2018,7(4): 167–186.

[4] Burch H B, Burman K D, Cooper D S. A 2011 survey of clinical practice patterns in the management of Graves’ disease [J]. J Clin Endocrinol Metab, 2012,97(12): 4549–4558.

[5] Bartalena L, Burch H B, Burman K D, et al. A 2013 European survey of clinical practice patterns in the management of Graves’ disease [J]. Clin Endocrinol (Oxf), 2016,84(1): 115–120.

[6] Negro R, Attanasio R, Grimaldi F, et al. A 2015 Italian Survey of Clinical Practice Patterns in the Management of Graves’ Disease: Comparison with European and North American Surveys [J]. Eur Thyroid J, 2016,5(2): 112–119.

[7] Momotani N, Hamada N, Ban Y. Management of thyrotoxicosis in women of childbearing age[J]. Folia Endocrinologica Japonica, 1977,53(10): 1148–1158.

[8] Jiang N, Li J, Zhang Z, et al. Preliminary evaluation of the pregnancy outcomes in women of reproductive age with graves’ hyperthyroidism after radioactive iodine therapy[J]. Thyroid, 2013,23: A51.

[9] Elston M S, Tu’akoi K, MEYER-Rochow G Y, et al. Pregnancy after definitive treatment for Graves’ disease--does treatment choice influence outcome?[J]. Aust N Z J Obstet Gynaecol, 2014,54(4): 317–321.

[10] Hawken C, Sarreau M, Bernardin M, et al. Management of Graves’ disease during pregnancy in the Poitou-Charentes Region[J]. Ann Endocrinol (Paris), 2016,77(5): 570–577.

[11] Zhang L H, Li J Y, Tian Q, et al. Follow-up and evaluation of the pregnancy outcome in women of reproductive age with Graves’ disease after 131Iodine treatment[J]. J Radiat Res, 2016,57(6): 702–708.

[12] Zhiwen Wu, Liyang Lai, Sheng Luo, et al. [the influence of thyroid function and pregnancy outcomes of women with hyperthyroidism became pregnancy after thyroidectomy] [J]. [Journal of Guangzhou medical university], 2018,46(02): 69–71.

[13] Mortimer R H, Cannell G R, Addison R S, et al. Methimazole and propylthiouracil equally cross the perfused human term placental lobule [J]. J Clin Endocrinol Metab, 1997,82(9): 3099–3102.

[14] Andersen S L, Olsen J, Wu C S, et al. Birth defects after early pregnancy use of antithyroid drugs: a Danish nationwide study[J]. J Clin Endocrinol Metab, 2013,98(11): 4373–4381.

[15] Honein M A, Kirby R S, Meyer R E, et al. The association between major birth defects and preterm birth[J]. Matern Child Health J, 2009,13(2): 164–175.

[16] Hamburger J I, Stoffer S S. Is Prevention of Hyperthyroidism Complicating Pregnancy Justification for Routine Ablative Therapy for Hyperthyroidism in Women in the Childbearing Years?[M]//Hamburger J I, Miller J M. Controversies in Clinical Thyroidology. New York, NY: Springer New York, 1981: 105–117.

[17] Kim Y A, Cho S W, Choi H S, et al. The Second Antithyroid Drug Treatment Is Effective in Relapsed Graves’ Disease Patients: A Median 11-Year Follow-Up Study[J]. Thyroid, 2017,27(4): 491–496.

[18] Sundaresh V, Brito J P, Wang Z, et al. Comparative effectiveness of therapies for Graves’ hyperthyroidism: a systematic review and network meta-analysis[J]. J Clin Endocrinol Metab, 2013,98(9): 3671–3677.

[19] Ren Z, Qin L, Wang J Q, et al. Comparative Efficacy of Four Treatments in Patients with Graves’ Disease: a Network Meta-analysis[J]. Exp Clin Endocrinol Diabetes, 2015,123(5): 317–322.

[20] Yuan J, Lu X, Yue Y. Comparison of curative effect of 131I and antithyroid drugs in Graves’ disease: a meta-analysis [J]. Minerva Endocrinol, 2018,43(4): 511–516.

[21] Burch H B, Cooper D S. ANNIVERSARY REVIEW: Antithyroid drug therapy: 70 years later[J]. Eur J Endocrinol, 2018,179(5): R261-R274.

[22] Laurberg P, Wallin G, Tallstedt L, et al. TSH-receptor autoimmunity in Graves’ disease after therapy with anti-thyroid drugs, surgery, or radioiodine: a 5-year prospective randomized study[J]. Eur J Endocrinol, 2008,158(1): 69–75.

[23] Polak M. Hyperthyroidism in early infancy: pathogenesis, clinical features and diagnosis with a focus on neonatal hyperthyroidism [J]. Thyroid, 1998,8(12): 1171–1177.

[24] Krude H, Biebermann H, Krohn H P, et al. Congenital hyperthyroidism[J]. Exp Clin Endocrinol Diabetes, 1997,105 Suppl 4: 6–11.

[25] Durham E, Howie R N, Parsons T, et al. Thyroxine Exposure Effects on the Cranial Base[J]. Calcif Tissue Int, 2017,101(3): 300–311.

[26] Ticconi C, Giuliani E, Veglia M, et al. Thyroid autoimmunity and recurrent miscarriage[J]. Am J Reprod Immunol, 2011,66(6): 452–459.

[27] Robertson J S, Gorman C A. Gonadal radiation dose and its genetic significance in radioiodine therapy of hyperthyroidism [J]. J Nucl Med, 1976,17(9): 826–835.

[28] Halnan K E. Radio-iodine treatment of hyperthyroidism-- a more liberal policy?[J]. Clin Endocrinol Metab, 1985,14(2): 467–489.

[29] Schneider D F, Sonderman P E, Jones M F, et al. Failure of radioactive iodine in the treatment of hyperthyroidism [J]. Ann Surg Oncol, 2014,21(13): 4174–4180.

[30] Laurberg P, Andersen S L. Therapy of endocrine disease: antithyroid drug use in early pregnancy and birth defects: time windows of relative safety and high risk?[J]. Eur J Endocrinol, 2014,171(1): R13-R20.

[31] Davis J R, Dackiw A P, Holt S A, et al. Rapid Relief: Thyroidectomy is a Quicker Cure than Radioactive Iodine Ablation (RAI) in Patients with Hyperthyroidism[J]. World J Surg, 2019,43(3): 812–817.

[32] Lin Y S, Lin J D, Hsu C C, et al. The long-term outcomes of thyroid function after subtotal thyroidectomy for Graves’ hyperthyroidism [J]. J Surg Res, 2017,220: 112–118.

[33] Shinall M J, Broome J T, Nookala R, et al. Total thyroidectomy for Graves’ disease: compliance with American Thyroid Association guidelines may not always be necessary[J]. Surgery, 2013,154(5): 1009–1015.

[34] Batra C M, Gupta V, Gupta N, et al. Fetal Hyperthyroidism: Intrauterine Treatment with Carbimazole in Two Siblings [J]. Indian journal of pediatrics, 2015,82(10): 962–964.

[35] Dierickx I, Decallonne B, Billen J, et al. Severe fetal and neonatal hyperthyroidism years after surgical treatment of maternal Graves’ disease [J]. Journal of Obstetrics and Gynaecology, 2014,34(2): 117–122.

[36] Laurberg P, Bournaud C, Karmisholt J, et al. Management of Graves’ hyperthyroidism in pregnancy: focus on both maternal and foetal thyroid function, and caution against surgical thyroidectomy in pregnancy[J]. European journal of endocrinology, 2009,160(1): 1–8.

